# Induction of labour during the COVID-19 pandemic: a national survey of impact on practice in the UK

**DOI:** 10.1101/2021.01.27.21250521

**Authors:** M Harkness, C Yuill, H Cheyne, SJ Stock, C McCourt, On behalf of the CHOICE Study Team

**Affiliations:** Nursing Midwifery and Allied Health Professionals Research Unit (NMHAP-RU), University of Stirling, Stirling, FK9 4NF; Centre for Maternal and Child Health Sciences, School of Health Sciences, City, University of London, 1 Myddleton Street, London, EC1R 1UB; Usher Institute, University of Edinburgh NINE Edinburgh BioQuarter, 9 Little France Road, Edinburgh EH16 4UX; MRC Centre for Reproductive Health, University of Edinburgh Queen’s Medical Research Institute, 47 Little France Crescent, Edinburgh EH16 4TJ

## Abstract

**Background:** Induction of labour (IOL) is one of the most commonly performed interventions in maternity care, with outpatient cervical ripening increasingly offered as an option for women undergoing IOL. The COVID-19 pandemic has changed the context of practice and the option of returning home for cervical ripening may now assume greater significance. This work aimed to examine whether and how the COVID-19 pandemic has changed practice around IOL in the UK.

**Method:** We used an online questionnaire to survey senior obstetricians and midwives at all 156 UK NHS Trusts and Boards that currently offer maternity services. Responses were analysed to produce descriptive statistics, with free text responses analysed using a conventional content analysis approach.

**Findings:** Responses were received from 92 of 156 UK Trusts and Boards, a 59% response rate. Many Trusts and Boards reported no change to their IOL practice, however 23% reported change in methods used for cervical ripening; 28% a change in criteria for home cervical ripening; 28% stated that more women were returning home during cervical ripening; and 24% noted changes to women’s response to recommendations for IOL. Much of the change was reported as happening in response to attempts to minimise hospital attendance and restrictions on birth partners accompanying women.

**Conclusions:** The pandemic has changed practice around induction of labour, although this varied significantly between NHS Trusts and Boards. There is a lack of formal evidence to support decision-making around outpatient cervical ripening: the basis on which changes were implemented and what evidence was used to inform decisions is not clear.

## Introduction

Induction of labour (IOL) is one of the most commonly performed interventions in maternity care, experienced by around 30.6% of pregnant women in the UK^1,2^. Cervical ripening is a key component of IOL^3^, whereby application of a drug or mechanical method over a number of hours causes softening, shortening, and opening of a woman’s cervix in preparation for labour. Cervical ripening is recommended by NICE for everyone undergoing IOL^4^.

Traditionally women undergoing cervical ripening are asked to remain in hospital throughout the procedure to allow maternity care staff to monitor their wellbeing and that of their baby. However, outpatient cervical ripening (commonly referred to as home cervical ripening) is increasingly being offered as an option, enabling women to choose to attend hospital for an initial assessment and administration of cervical ripening agent; and then to return home for a period of time (usually 24 hours), before returning to hospital to be reassessed. There is currently a lack of evidence to guide decision-making about where cervical ripening should happen^5^. The CHOICE Study, a UK-wide NIHR-funded prospective cohort study and process evaluation, aims to determine whether outpatient cervical ripening during induction of labour is safe, effective and cost effective^6^. A qualitative component of the CHOICE Study, qCHOICE, will also consider the acceptability of home cervical ripening to women and their families.

In March 2020, the World Health Organisation (WHO) declared the novel coronavirus (COVID-19) outbreak a global pandemic. Subsequently, in an attempt to control the virus, the UK government imposed national restrictions on movement and social contact: across the National Health Service (NHS) this changed how, when and where people received their maternity care^7^. For women this has resulted in reduced choice about place of birth and restrictions on birth partners accompanying them when they attend hospital, even during labour and birth^8^. Within the context of these sudden, substantial and often distressing changes to service provision, the offer to return home after cervical ripening may have assumed new significance for women and their health care professionals when making decisions about childbirth.

The CHOICE Study commenced in early 2020, and changes to practice in response to the pandemic inevitably alter the context in which the study will be conducted. It is important to understand changes in policy and practice surrounding IOL to inform the CHOICE study, and other research, and to understand how health care professionals can best support women who are making decisions about their induction of labour. Consequently, the CHOICE Study team conducted a survey of UK maternity services in order to establish whether and how the pandemic has changed practice around induction of labour in the UK, particularly in relation to home and hospital cervical ripening.

## Methods

### Data Collection

This survey aimed to determine whether aspects of practice and policy had altered in response to the pandemic. The research team carefully considered the process of IOL, with a focus on cervical ripening, and key elements of that process formed the basis of a questionnaire. The questionnaire comprised fixed response questions, with free-text boxes requesting additional detail, allowing respondents to provide written comments about key issues.

Women’s experience of the IOL process is vital, and although it was beyond the scope of this work to ask their views directly, the survey did endeavour to determine perceived changes in women’s response to IOL.

All NHS Trusts and Boards in the UK that offer maternity services were identified through the NHS service directories for the four UK countries (n=157). Senior midwifery and obstetric staff at those Trusts and Boards were contacted by email through established networks including the RCM Heads of Midwifery network, professional contacts of CHOICE Study team; local Clinical Research Networks; and the British Intrapartum Care Society. The questionnaire was hosted by *Online Surveys* (www.onlinesurveys.ac.uk), and a Microsoft Word version was available should the respondent have difficulties accessing the online survey site. The survey was open between June 2020 and November 2020.

### Data Analysis

Responses were identified according to NHS Trusts and Boards, as opposed to individual maternity units, and where duplicate responses were received those were compared and one combined response created. All UK Trusts and Boards currently offering maternity services (n=156) was used as the denominator to calculate response rate.

Responses were exported from the survey site into IBM SPSS Statistics 23 software and descriptive statistics were produced in relation to the key issues. Free-text responses were analysed using a conventional content analysis approach to determine what themes are present and how often they reoccur in the data^9^. This approach was employed, as it is more phenomenological in nature, best suited for research on a topic where there is limited pre-existing literature. Preconceived codes and categories are not used; they instead ‘flow’ from the data. Each line of free-text responses was coded, and the resulting codes were grouped into categories and sub-categories, creating “meaningful clusters” (9, p1279). In order to establish the validity and credibility of the content analysis, emergent categories were triangulated with the descriptive statistics outcomes.

## Ethical Approval

Ethics approval was obtained from the York & Humber – Sheffield Research Ethics Committee in June 2020 (IRAS: 276788) as part of the wider CHOICE Study application. The survey was initiated in response to Committee questions about the impact of COVID-19 on IOL during the Proportionate Review.

## Findings

One hundred and fifty-seven UK NHS Trusts and Boards that currently offer maternity services were identified. One maternity unit had suspended all services due to the pandemic making 156 Trusts and Boards eligible for inclusion. Completed responses were received from 92 of the 156 (59%). A breakdown of response by UK countries is shown in Table 1. The survey was most often completed by a Senior Midwife or a Consultant Obstetrician (Table 2).

**Table 1:**
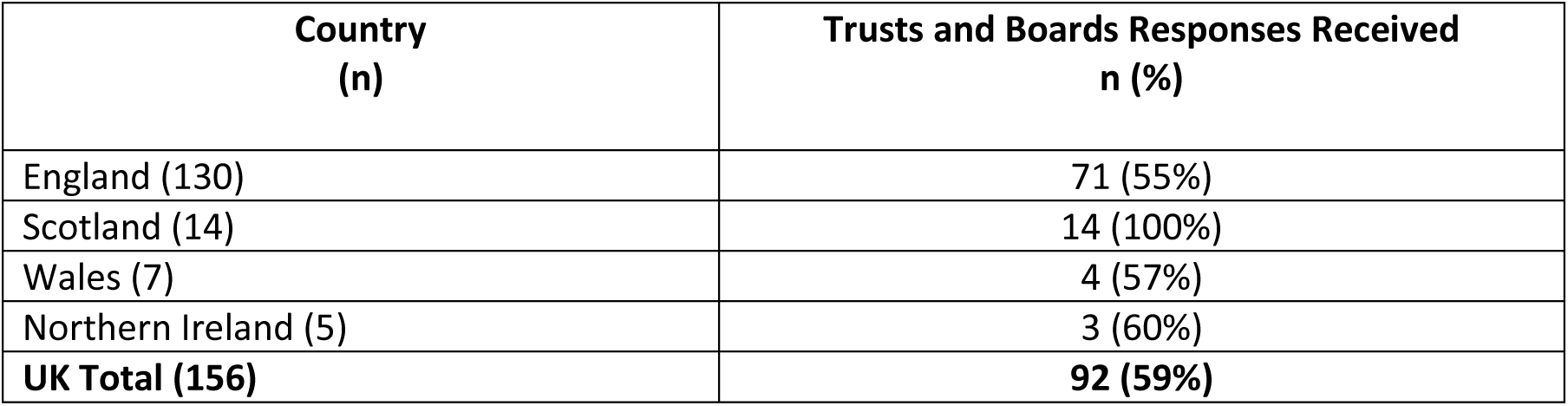
Response rate by UK Country

| **Country**<br>**(n)** | **Trusts and Boards Responses Received**<br>**n (%)** |
| --- | --- |
| England (130) | 71 (55%) |
| Scotland (14) | 14 (100%) |
| Wales (7) | 4 (57%) |
| Northern Ireland (5) | 3 (60%) |
| **UK Total (156)** | **92 (59%)** |

**Table 2:** Work role of person who completed the survey

| **Work role of person completing the survey** | **n** |
| --- | --- |
| Senior Specialist Midwife or Midwifery Manager | 31 |
| Consultant Obstetrician | 25 |
| Head of Midwifery | 24 |
| Consultant Midwife | 16 |
| Research Midwife | 8 |
| Clinical Director | 4 |
| Clinical Midwife | 3 |
| Obstetric Trainee (ST7) | 1 |
| **Total** | **112\*** |

\*Some responses stated that two or more people had completed the survey together

Two hundred and fifty-five unique, free-text responses were collected across 11 survey questions. Three key categories were identified from the content analysis: IOL practice, Changes to IOL practice and Changes in IOL uptake, within which there were several sub-categories (Tables 3, 6 & 7).

**Table 3:** Outcomes from free-text response content analysis -IOL Practice

| Category | Sub-category | Examples from data |
| --- | --- | --- |
| IOL practice | OP IOL eligibility was limited | "Low risk post-dates induction living within 20 minutes drive of the hospital."<br>"Post-dates (40+12 days), Midwifery led care pathway Age between 18-40 BMI < 35 Lives within 1 hour of hospital Singleton pregnancy No medical or obstetric history/ complications No more than 4 previous births"<br>"In order to be suitable for outpatient IOL, the woman must meet the following criteria: under midwifery-led care during the pregnancy; aged 18-39; primigravid; BMI 18-34; gestational age of 41+1 - 41+6"<br>"Low risk T+10-14; live within 30 minutes of the hospital and have their own transport. Induction for post dates only with normal liquor volume and otherwise low risk."<br>"Multips who live within 30mins of the consultant unit with no history of reduced FM & low risk. Low risk post-dates primips" |
|  | Midwife-led IOL | "inserted by the midwife after obstetric review. In case of difficulty, the obstetricians get involved."<br>"Midwives do the in-patients, doctors currently do the out-patient inductions -but it is becoming more accepted and I think that will change."<br>"Obstetricians insert Dilapan currently. "Midwives insert Prostin/Propress." |

### IOL practice

The mean reported induction rate across all responding Trusts and Boards was 34%. Figure 1 shows the reported rate in the past year across all responding units. The mean date at which induction of labour was routinely offered for reason of gestational length was ten days after term (40weeks +10days).

**Figure 1:**
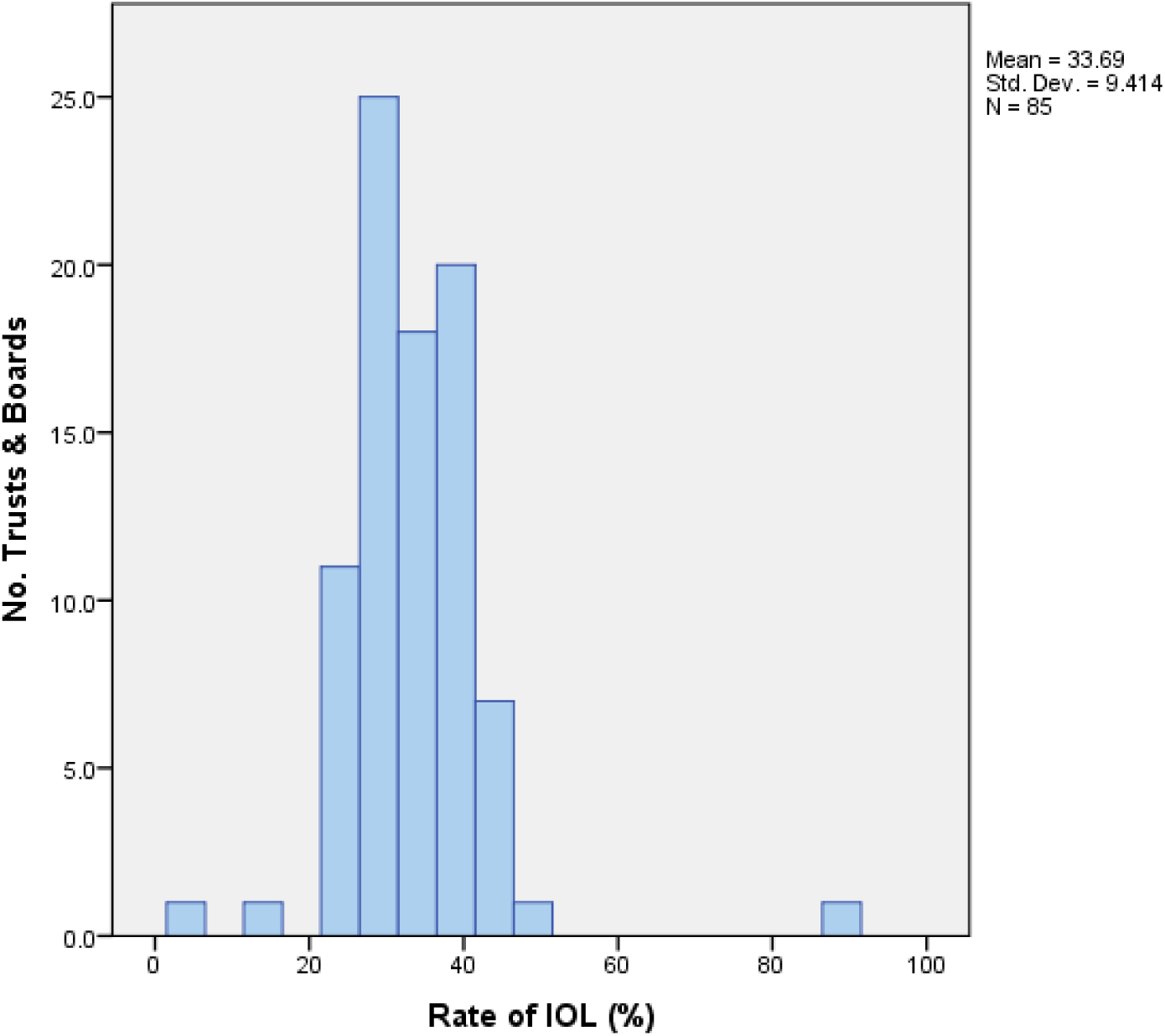
Reported rate of IOL per unit in the past year.

Ninety (98%) Trusts and Boards reported using pharmaceutical methods, such as Propess (PGE2), for cervical ripening, and 64 (70%) mechanical methods, including Cooks balloon and Dilapan-S. At most Trusts and Boards, 74 (80%), both midwives and obstetricians conducted cervical ripening. This collaboration was reflected in the free-text responses, although respondents reported a generally midwifery-led practice (Table 3).

### Changes in IOL practice due to COVID-19

A majority of Trusts and Boards reported no change to their IOL rates and practice in response to the pandemic. For those that did, the survey determined significant change to key aspects of IOL practice (Tables 4 & 5).

**Table 4:**
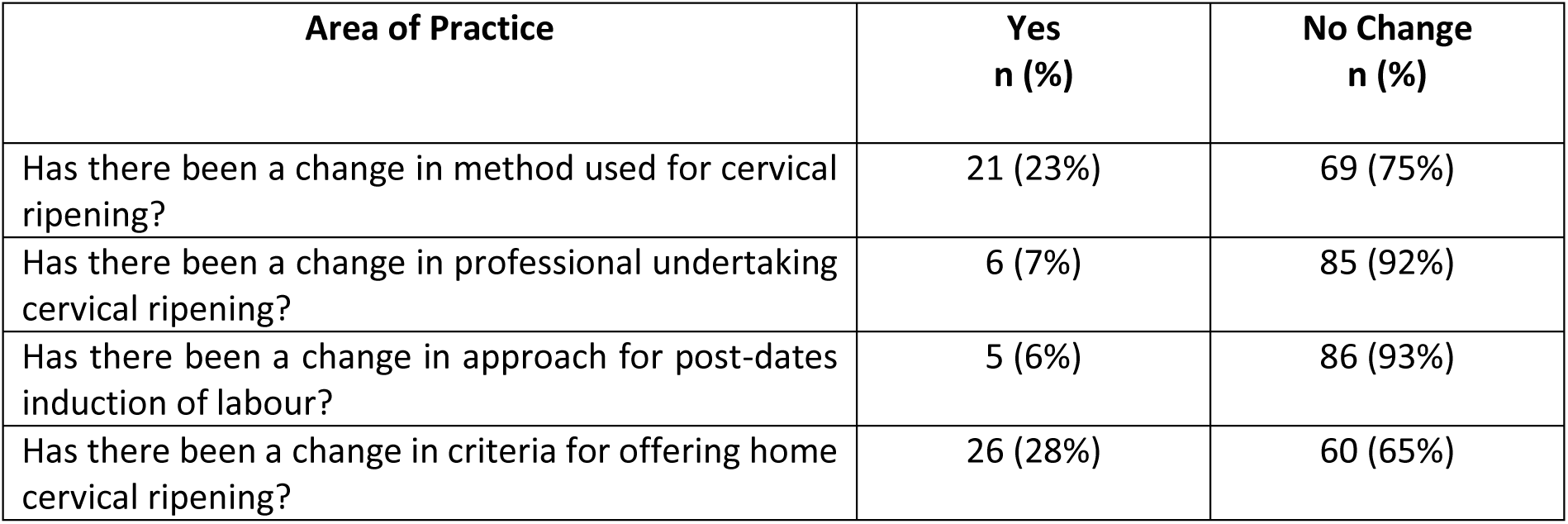
Significant change to key aspects of IOL practice

| **Area of Practice** | **Yes**<br>**n (%)** | **No Change**<br>**n (%)** |
| --- | --- | --- |
| Has there been a change in method used for cervical ripening? | 21 (23%) | 69 (75%) |
| Has there been a change in professional undertaking cervical ripening? | 6 (7%) | 85 (92%) |
| Has there been a change in approach for post-dates induction of labour? | 5 (6%) | 86 (93%) |
| Has there been a change in criteria for offering home cervical ripening? | 26 (28%) | 60 (65%) |

**Table 5:** Change in number of women returning home after starting cervical ripening in response to the pandemic

| **More women**<br>**discharged home after**<br>**cervical ripening** | **Fewer women**<br>**discharged home after**<br>**cervical ripening** | **No change** | **Missing** |
| --- | --- | --- | --- |
| 26 (28%) | 5 (5%) | 56 (61%) | 7 (%) |

Twenty-one (23%) Trusts and Boards stated the methods used for cervical ripening had changed in response to the pandemic, while 69 (75%) were unchanged. We asked respondents to provide details of changes: most often they reported a switch to mechanical methods, but there was little information provided about why that was the case. Switching to use of Dilapan-S as a method of cervical ripening was notable, and often was mentioned in relation to participation in the SOLVE trial (ISRCTN20131893) (Table 6).

**Table 6:** Outcomes from free-text response content analysis -changes to IOL practice

| Category | Sub-category | Examples from data |
| --- | --- | --- |
| **Changes to IOL practice** | **More OP IOL offered** | “Pregnant women deemed low risk are offered out patient induction. Criteria for induction changed. Softer reasons such as Social / maternal age are reviewed.” |
|  | **Changes to IOL methods and process** | “We brought forward the mechanical ripening and ALL women who had Dilapan were offered home. Dilapan introduced, expectation that women will go home during Dilapan ripening. “<br>“We are very keen to offer birth in a FMLU following OIOL but do not have enough data locally to support this until OIOL births numbers increase on our AMLUs” |
|  | **Increased OB presence** | “Initially only obstetricians trained to use balloon catheter. Midwives are now being trained to conduct balloon catheter IOL”<br>“Doctors still doing Dilapan. Midwives mostly doing prostaglandins. More obstetric involvement as Dilapan training not yet rolled out to midwives.” |

Eighty-five (92%) reported no change in the professional undertaking cervical ripening, six units (7%) did report change, and one response was missing. Within the free-text responses explaining changes, the six respondents most often noted that obstetricians had taken over responsibility for the procedure, and this appeared to be linked to change in the method being used, with obstetricians receiving training for the change ahead of midwives (Table 6).

Though 86 (93%) Trusts and Boards stated no change in approach regarding IOL for gestational age in response to the pandemic, the services that did change revealed sometimes dramatic shifts in practice. For instance, one island maternity unit noted increased reluctance among women to travel to the tertiary unit on the mainland during the pandemic. This meant that people normally considered to be at higher risk, for example those with a raised BMI or of age 40 and older, were now offered postdates induction of labour at the island obstetric unit instead of being flown to the mainland.

### Impact on home cervical ripening

Prior to the pandemic, 50 (54%) of Trusts and Boards offered women the option to return home after insertion of pessary or device for cervical ripening, while 42 (46%) did not offer the option. Thirty nine of the 50 (78%) respondents from units that offered home cervical ripening stated that only certain categories of women are offered this option. Being categorised as ‘low risk’ was the most frequently mentioned criteria for offering women this choice.

The stated criteria for offering home induction varied across the Trusts and Boards, and in some cases, suggested that eligibility was limited (Table 3). The most common criteria included post-dates, maternal choice, absence of clinical indicators of risk (e.g. history of reduced fetal movements, increased BMI, maternal age over 40 years, maternal medical conditions, fetal medical concerns). The option was also reportedly offered to women who had balloon catheter, were on a midwifery-led pathway or planned to give birth in a midwifery unit. Other responses noted that women’s personal circumstances also influenced the availability of this option for women. Personal criteria for eligibility included living with 20-60 minutes by car from hospital, having access to a phone or own transport, having another adult present at home and being able to communicate effectively with staff.

Twenty-six (28%) Trusts and Boards reported that criteria for offering home IOL had changed in response to the pandemic, 60 (65%) that it had not changed. There were six missing responses. Overall, respondents reported that more women are being discharged home for cervical ripening (Table 5). Comments were more likely to describe a shift towards offering home cervical ripening, often linked to a change in method from pharmaceutical to mechanical. When a change in policy for offering home cervical ripening had been in progress prior to the pandemic, some respondents reported the process of introduction being either accelerated or halted. A small number noted that they had stopped offering women the opportunity to return home after commencing cervical ripening, one stating that this was in an effort to reduce multiple attendance at hospital by patients and their visitors.

### Women’s responses to IOL as reported by staff

Twenty-two (24%) of respondents noted a change in women’s responses to recommendations for IOL in relation to gestational age in light of the pandemic. Seventy (76%), had not noted a change. Free-text responses indicated that changes in IOL uptake were mostly related to women’s reluctance to the procedure, partner restrictions and desire to stay home or local (Table 7).

**Table 7:**
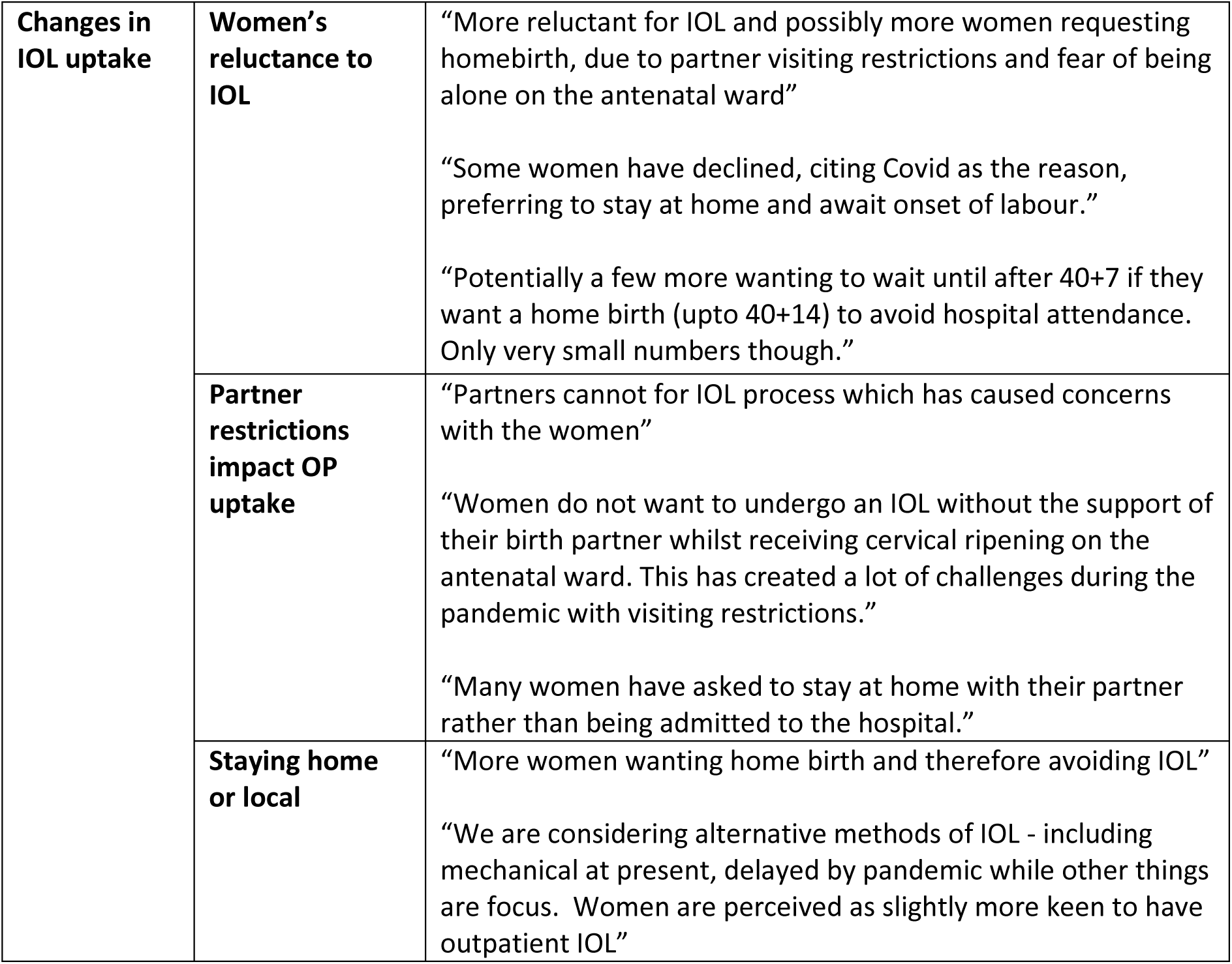
Outcomes of free-text response content analysis -changes in IOL uptake

|  |  |  |
| --- | --- | --- |
| **Changes in IOL uptake** | **Women's reluctance to IOL** | "More reluctant for IOL and possibly more women requesting homebirth, due to partner visiting restrictions and fear of being alone on the antenatal ward"<br>"Some women have declined, citing Covid as the reason, preferring to stay at home and await onset of labour."<br>"Potentially a few more wanting to wait until after 40+7 if they want a home birth (upto 40+14) to avoid hospital attendance. Only very small numbers though." |
|  | **Partner restrictions impact OP uptake** | "Partners cannot for IOL process which has caused concerns with the women"<br>"Women do not want to undergo an IOL without the support of their birth partner whilst receiving cervical ripening on the antenatal ward. This has created a lot of challenges during the pandemic with visiting restrictions."<br>"Many women have asked to stay at home with their partner rather than being admitted to the hospital." |
|  | **Staying home or local** | "More women wanting home birth and therefore avoiding IOL"<br>"We are considering alternative methods of IOL - including mechanical at present, delayed by pandemic while other things are focus. Women are perceived as slightly more keen to have outpatient IOL" |

The responses suggest that women’s reluctance to IOL was related to less uptake or delay of the procedure, and that decisions were interlinked with restrictions on birth partners accompanying them during a hospital stay. Interestingly, it was more often reported that women opted to decline or delay induction of labour, rather than opt for home cervical ripening. There were further reports of women requesting elective caesarean section rather than IOL and of fewer requests for IOL before clinical indications. Declined or delayed IOL was also connected, by respondents, to local increases in planned home birth rates.

The survey also asked if there had been a change in women’s response to offer of home cervical ripening in relation to the pandemic. There were 20 missing responses to this question; of those who provided an answer, 23 (32%) respondents reported that more women wanted home IOL, two that fewer women wanted outpatient IOL, and 47 (65%) that there had been no change. Comments around this again indicate that women preferred to delay or avoid IOL altogether, and that the shift to outpatient IOL was clinician-driven rather than service user-driven.

## Discussion

The survey was completed by senior midwives and obstetricians at a large proportion of UK NHS Trusts and Boards and provides important insight to current practice surrounding induction of labour, as well as how that has changed in response to the COVID-19 pandemic.

We found that over half of NHS Trusts and Boards offered home cervical ripening prior to the COVID-19 pandemic, that 28% had changed their criteria for offering home cervical ripening in response to the pandemic, and that almost a third now report more women going home after having this procedure. This shift towards home cervical ripening was accompanied by an apparent move towards using mechanical methods, such as balloon catheter and Dilapan-S, which were often considered safer and more acceptable for use at home than more established pharmaceutical methods. Conversely, some Trusts and Boards withdrew their home cervical ripening service.

Although the survey found significant changes in practice at some Trusts and Boards, this was not perhaps as great as might be expected given the context of the substantial changes imposed to service provision in order to limit physical contact at national level. The diversity of responses between Trusts and Boards may be due in part to varying geographies and populations as well as different experiences of the pandemic. However, the survey respondents often linked policy decision-making to those restrictions that were imposed across NHS services in response to the pandemic, particularly the impetus to minimise hospital admission and length of stay, and restrictions on birth partners accompanying women to hospital. At present there is little evidence to support home versus hospital cervical ripening or the methods used to achieve it^5^ and decision-making around changes to practice seem likely to have been driven by immediate need to reduce physical contact in response to the pandemic. Recent professional scrutiny and reflection recognises that some changes that were made were unnecessary and potentially harmful, and revised guidance concerning service level response to the pandemic has now been issued^11^. It is not clear whether the changes in policy and practice found in this survey are permanent or whether they will be subject to further scrutiny, either in response to guidance from obstetric and midwifery professional bodies or when COVID-19 is no longer a factor in service provision.

Interestingly, some respondents reported an impression that the pandemic had improved efficacy of clinical decision-making and support among clinicians for change, and this seems to be corroborated by the significant amount of change in policy observed to have happened in a relatively short space of time. The changes made during this short period were substantial and varied: introducing home cervical ripening where it had not previously been considered; accelerating introduction when a service had been under consideration; or stopping an already running home cervical ripening service. These are complex and multi-faceted changes to policy and practice, demonstrating that changes which often take considerable time to implement can be fast-tracked when there is an urgent need to do so. However, it does not provide insight to the decision-making process or to the quality of decisions that were made.

Respondents often mentioned external factors as influencing service provision. Participation in the SOLVE trial (a randomised control trial comparing Dilapan-S rods with Propess pessary) was frequently mentioned as the driver for introduction of a home induction service, and conversely pausing the SOLVE trial in response to the pandemic led to withdrawal of home cervical ripening service in some areas. Staff training, particularly in use of mechanical methods of cervical ripening, also influenced whether a service was made available. In most Trusts and Boards both obstetricians and midwives conducted cervical ripening, but many noted that either only obstetricians or only midwives had received training in insertion of balloon catheters or Dilapan-S, and that this affected whether home cervical ripening was available to women. Inconsistency in this area may reflect lack of evidence about the use of mechanical methods, ultimately creating difference in the options available to women depending on where they access care.

Women’s response to induction of labour is also reported to have changed, and this was most often linked to restrictions on birth partners accompanying them to hospital. Women were observed as adapting their decision-making, most notably by choosing alternatives to induction of labour that might reduce the amount of time they spend in hospital. In some cases, it appears that women’s power to choose was increased: as criteria for induction of labour changed options that were previously deemed unsafe or lacking in evidence became acceptable, including women who can now opt for induction at their home island obstetric unit, or those who now have the option to return home for cervical ripening when they could not before. However, it also appears that for some women decisions were primarily motivated by fear of being alone during labour and birth.

In some Trusts and Boards, the option of home cervical ripening was unavailable to women because the service was withdrawn, and when it was offered ability to choose this option was severely limited through imposition of eligibility criteria. It is notable that the women most likely to be denied the option of home cervical ripening are those who may already feel marginalised within maternity services: women categorised as ‘high risk’; women affected by poverty; women whose first language is not English; and women with disabilities that alter their ability to communicate. Current evidence on women’s experiences of home IOL is limited^12^, but the research to date indicates its potential benefits for women as compared with the in-hospital experience^13^. Given the general paucity of research on home IOL, the qCHOICE study (NIHR127569) will explore women’s, partners’ and healthcare professionals’ experiences and perceptions of home cervical ripening in more depth.

## Conclusion

It is clear from this research that the pandemic has changed practice around IOL, although the response varied significantly between NHS Trusts and Boards. There is a lack of formal evidence to support decision-making around home cervical ripening, and the basis on which changes were implemented and what evidence was used to inform decisions is not clear. This affirms disparity and inequity of choice for women who are offered IOL, dependent on the areas in which they live and access maternity services.

Further research, such as the CHOICE Study, is crucial for providing evidence to inform and support decision-making by women and their care givers. In the future, when COVID-19 is no longer a factor in clinical decision-making, changes to practice made during the pandemic should be scrutinised within the context of this new evidence.

## Data Availability

The data sets used and/or analysed during the current study are available from the corresponding author on reasonable request.

## List of Abbreviations

COVID & COVID-19: The name given by the World Health Organization (WHO) for the disease caused by the novel coronavirus SARS-CoV2.
AMLU: Alongside Midwifery Led Unit
BMI: Body Mass Index
CHOICE: The CHOICE Study
FM: Fetal Movements
FMLU: Freestanding Midwifery Led Unit
IOL: Induction of Labour
NICE: National Institute of Clinical Excellence
NHS: National Health Service
OIOL: Outpatient Induction of Labour
qCHOICE: The qCHOICE Study
WHO: The World Health Organisation
UK: United Kingdom

## Declarations

### Competing Interests

The authors declare they have no competing interests.

### Funding

CHOICE is funded by the National Institute of Healthcare Research Health Technology and Assessment (NIHR HTA) NIHR 127569. The views expressed are those of the authors and not necessarily those of the NIHR or the Department of Health and Social Care.

### Author Contributions

Dr Mairi Harkness: Created draft of work; Substantial contribution to acquisition, analysis and interpretation of data; Substantially revised the work; Approved submitted version

Dr Cassandra Yuill: Substantial contribution to conception and design of the work; Substantial contribution to analysis and interpretation of data; Substantially revised the work; Approved submitted version.

Professor Helen Cheyne: Substantial contribution to conception and design of the work; Substantial contribution to interpretation of data; Substantially revised the work; Approved submitted version.

Professor Christine McCourt: Substantial contribution to conception and design of the work; Substantial contribution to interpretation of data; Substantially revised the work; Approved submitted version.

Dr Sarah Stock: Substantial contribution to conception and design of the work; Approved submitted version.

## Acknowledgements

Thanks are due to the staff who completed the survey.

The authors would also like to thank the CHOICE Study Team:

Dr Amarnath Bhide, Consultant Obstetrician, St George’s University Hospitals Trust; Dr Mairead Black, Senior Clinical Lecturer, University of Aberdeen; Mrs Maggie Reid, Research Midwife, University of Edinburgh; Mrs Fiona Wee, Trial Manager, University of Edinburgh; Miss Dikshyanta Rana, Research Assistant, University of Glasgow; Dr Kathleen Boyd, Senior Lecturer, University of Glasgow; Professor Julia Sanders, Professor of Clinical Nursing & Midwifery, Cardiff University; Miss Neelam Heera, Patient and Public Involvement Representative; Mrs Jane Huddleston, Patient and Public Involvement Representative; Professor Fiona Denison, Reader/Honorary Consultant in Maternal and Fetal Medicine, University of Edinburgh; Professor Dharmintra Pasupathy, Professor of Maternal Fetal Medicine, University of Sydney; Professor Neena Modi, Professor of Neonatal Medicine, Imperial College London; Professor Gordon C S Smith, Professor of Obstetrics and Gynaecology, The University of Cambridge; Professor John Norrie, Professor of Medical Statistics and Trial Methodology / Director of Edinburgh Clinical Trials Unit / Co-Head of Centre for Population Health Sciences, University of Edinburgh

## Notes

### Competing Interest Statement

The authors report research grants paid to institutions from: 
NIHR; Wellcome Trust; Tommy's Charity, MRC
during the course of the work

### Author Declarations

Ethics approval was obtained from the York & Humber Sheffield Research Ethics Committee in June 2020 (IRAS: 276788) as part of the wider CHOICE Study application. The survey was initiated in response to Committee questions about the impact of COVID-19 on IOL during the Proportionate Review.

